# Beyond Acute Jaundice: refining case definitions for suspected hepatitis E in South Sudan and Bangladesh

**DOI:** 10.1101/2025.10.03.25337273

**Authors:** Aybüke Koyuncu, Repon C. Paul, Robin Nesbitt, Kinya Vincent Asilaza, Catia Alvarez, Kajal C. Banik, Joseph Wamala, Arifa Nazneen, Melat Haile, Shariful Amin Sumon, Etienne Gignoux, Kishor K. Paul, Manuel Albela, Arifa Akram, Frederick Beden Loro, M. Salim Uzzaman, Duol Biem, Monica Rull, Stephen P. Luby, Isabella Eckerle, John Rumunu, Iza Ciglenecki, Emily S. Gurley, Andrew S Azman

## Abstract

The true burden of hepatitis E is poorly understood due to a lack of routine diagnostic testing and nonspecific symptoms. We evaluated the sensitivity and specificity of international case definitions for suspected hepatitis E and explored whether alternative case definitions could more accurately identify those with true hepatitis E, thus improving our ability to interpret surveillance data. We used data from acute jaundice surveillance in South Sudan (March-December 2022) and Bangladesh (December 2014-September 2017), two settings where hepatitis E genotype 1 circulates. Individuals seeking care with acute jaundice syndrome (AJS) were asked about signs/symptoms of hepatitis E and underwent testing with anti-hepatitis E virus (HEV) IgM ELISA. We used an ensemble of classification models to assess how well signs/symptoms could distinguish between AJS with and without HEV. To explore alternative case definitions, we estimated the sensitivity and specificity of all combinations of signs/symptoms. Among patients with AJS, 20% in South Sudan and 38% in Bangladesh had IgM antibodies. International case definitions for suspected hepatitis E had variable sensitivity (53-96%) and specificity (6-60%) across study populations. Alternative case definitions had poor discrimination in both populations (AUC_SouthSudan_=0.64; 95% Confidence Interval (CI): 0.57, 0.71; AUC_Bangladesh_= 0.60; 95% CI: 0.57, 0.63). No alternative case definitions had both sensitivity and specificity above 60% in both study populations. Acute jaundice surveillance in two distinct populations revealed that signs/symptoms alone are insufficient to distinguish HEV-related cases from other causes of acute jaundice. Accurately characterizing the burden of hepatitis E and assessing the cost effectiveness of vaccines will require expanded use of diagnostic testing.

## Introduction

Hepatitis E genotypes 1 and 2 disproportionately affect populations in low-income settings with limited resources for surveillance and outbreak response (1). Hepatitis E virus (HEV) is an epidemic-prone pathogen transmitted through the fecal-oral route, and universal access to clean water, sanitation, and hygiene (WASH) is needed for long-term control and elimination (1). The risk of severe outcomes due to HEV infection is heightened in pregnant women, for whom case fatality risks can be up to 25% and the odds of intrauterine fetal demise are more than double compared to uninfected women (2–5).

HEV239 vaccine (Hecolin; Innovax, Xiamen, China) is safe and highly efficacious in preventing hepatitis E disease (6,7), but the World Health Organization (WHO) has not recommended the routine use of the vaccine in HEV-endemic countries, citing insufficient data on the incidence of hepatitis E in the general population as a key barrier (8). Understanding the burden of hepatitis E is complicated by a lack of routine testing for hepatitis E among acute jaundice cases, and non-specific symptoms (e.g., acute jaundice, fever) that are shared with other diseases (9). Though diagnostics for hepatitis E can have high sensitivity and specificity (10–13), many settings where hepatitis E occurs lack the laboratory infrastructure needed for confirmatory testing of suspected hepatitis E cases and also limited availability of rapid diagnostic tests that can be used at point-of-care. Though context-dependent, the cost of RDTs (4-11.5 USD per test (14)) is likely to remain a barrier to broad use in the near future, especially since hepatitis E treatment is typically supportive care (3) and diagnostic confirmation typically does not change patient management. In the absence of diagnostic capacity, surveillance for hepatitis E in low-income settings is typically based on diagnosis of acute jaundice syndrome (AJS) (15), and outbreaks are typically identified when the number of AJS cases that seek care at a facility is higher than expected during a defined period or if clusters of fulminant hepatitis and deaths are identified in pregnant women. Reports of hepatitis E outbreaks in the published and grey literature frequently cite uncertain and/or incorrect initial conclusions about outbreak aetiology, and outbreaks are often initially suspected to be due to malaria (16) or yellow fever (17,18). A better understanding of the likelihood that acutely jaundiced individuals truly have hepatitis E can help decision-makers understand when pathogen-specific outbreak response activities are needed.

The WHO hepatitis E outbreak toolbox includes several international case definitions for suspected hepatitis E proposed by WHO and other non-governmental organizations (NGOs) (19). The suspected case definitions include different combinations of signs and symptoms that correspond to acute illness (e.g., fever) and signs of liver damage (e.g., jaundice, nausea), and existing guidelines do not distinguish between epidemiologic contexts. Like many clinical case definitions, those for hepatitis E are known to have poor specificity however no published studies to date have quantified the performance of these case definitions. Furthermore, it remains unclear to what extent the inclusion of symptoms other than acute jaundice syndrome improves specificity and whether any signs/symptoms, even when used in a more complex manner, can consistently distinguish AJS with and without HEV across settings. More precise quantification of existing case definitions and exploration of potentially more informative ones, may allow for modest improvements in guidelines for hepatitis E surveillance in low resource settings, and allow for estimates of the “true” burden of disease from imperfect surveillance data.

Here, we leverage data from facility-based acute jaundice surveillance in South Sudan (predominantly outpatients) and Bangladesh (inpatients), two settings where HEV genotype 1 transmission circulates, to evaluate and compare the sensitivity and specificity of international case definitions for identifying suspected hepatitis E compared to laboratory-confirmed hepatitis E and explore if there are alternative case definitions with improved specificity.

## Materials & Methods

### Surveillance site: South Sudan

Hepatitis E cases have been documented in Bentiu internally displaced persons (IDP) camp in South Sudan since 2014 with large outbreaks in 2015 (2189 reported cases) and 2016 (924 reported cases) (7). In response to an outbreak in March 2022, the South Sudan Ministry of Health, in partnership with Médecins Sans Frontières (MSF), implemented a reactive vaccination campaign against hepatitis E (HEV239; Hecolin; Innovax, Xiamen, China) in the camp (21), coupled with a vaccine effectiveness study. As part of the study, MSF conducted comprehensive diagnostic testing for all patients with AJS identified at the MSF facility located in the camp from March to December 2022 (7). AJS was defined as the acute (recent, new, or sudden) onset of yellow coloration of the whites of the eyes or skin, dark urine, or pale clay stools. All cases of AJS were considered suspected cases of hepatitis E.

All suspected hepatitis E cases seeking care at the MSF hospital (outpatient and inpatient departments) were identified by clinicians and referred to the study team after consultation or hospital admission. Study staff explained the study objectives and, among suspected cases willing to participate, obtained consent for participation. Adults provided written informed consent, while individuals under 18 years of age provided assent, and their guardians provided written informed consent. Study staff administered a questionnaire on demographics, vaccination status, and signs and symptoms including symptoms of AJS. A laboratory technician collected a venous blood sample and prepared all specimens for testing, storage, and transport. Details on specimen storage and handling have been described elsewhere (7).

### Laboratory methods: South Sudan

Testing for IgM ELISA was conducted in a reference laboratory at the University Hospitals of Geneva. We used WANTAI HEV-IgM ELISA (WE-7196, Beijing Wantai Biological Pharmacy Ent.) to detect HEV antibodies in venous plasma following the package insert. Samples with specimen absorbance values (A) greater than or equal to the cut-off-value (C.O) (A/C.O≥1) were considered to have detectable IgM. All samples were also tested for the presence of malaria histidine-rich protein 2 (a marker of acute malaria infection) using a Paracheck rapid diagnostic test (Orchid Biomedical Systems, Goa, India), hepatitis A infection based on the detection of HAV RNA using AltoStar HAV RT-PCR assay, (Altona Diagnostics, Hamburg, Germany), hepatitis B infection based on the detection of hepatitis B surface antigen (a marker of acute or chronic hepatitis B infection) using a SD Bioline rapid diagnostic test (Abbott, Orlando, FL, USA), and hepatitis C infection based on the detection of anti-hepatitis C virus antibodies using a SD Bioline rapid diagnostic test (Abbott, Orlando, FL, USA).

### Surveillance site: Bangladesh

Hepatitis E transmission in Bangladesh is endemic with large seasonal outbreaks. Surveillance methods in Bangladesh have been previously described (4). From December 2014 to March 2015 the International Centre for Diarrhoeal Disease Research, Bangladesh (icddr,b) supported hospital-based acute jaundice surveillance in six tertiary hospitals (five government teaching hospitals and one private teaching hospital) located in five of the seven divisions in Bangladesh. Tertiary hospitals in Bangladesh generally serve severely ill patients, some of whom have been referred from lower-level hospitals in districts and sub-districts near the tertiary hospital. In each hospital, a physician working with a field assistant from the International Centre for Diarrhoeal Disease Research, Bangladesh (icddr,b) reviewed daily admission records in the obstetrics and gynecology ward, adult medicine ward, and intensive care unit to identify AJS in-patients at least 14 years of age. AJS was defined as a new onset of either yellow eyes or skin within the past 3 months and continuing on the day of admission.

The physician in each hospital approached all eligible patients and sought consent from the patient, or their guardian in the case of severely ill patients, for study enrollment. Patients over 17 years of age provided written informed consent, while patients 14 to 17 years of age provided written assent, and their guardian provided written informed consent. The physician recorded each consenting patient’s illness history and relevant clinical information, completed a questionnaire on patient demographics, collected a 5 mL blood specimen for laboratory testing, and monitored patient vital status for the duration of hospitalization. The physician asked patients whether symptoms were present at any time during their illness and whether the symptom was continuing on the day of admission. Among patients who were discharged, field assistants called all enrolled patients 3 months post-discharge to ascertain vital status. If patients did not have a phone, field assistants visited their home for follow-up. Post-discharge follow-up was not possible for patients admitted after June 30, 2017, due to the end of the surveillance period on September 30, 2017.

### Laboratory methods: Bangladesh

Field assistants centrifuged blood specimens on the day of sample collection and stored serum at the hospital in a liquid nitrogen dry shipper. Samples were transported to a laboratory in Dhaka every two weeks. Once at the laboratory in Dhaka, samples were kept in a freezer at - 80°C until testing. Samples were tested using WANTAI HEV-IgM ELISA (WE-7196, Beijing Wantai Biological Pharmacy Ent.; A/C.O≥1 considered detectable IgM) to detect HEV antibodies in serum following the package insert. All samples were also tested for hepatitis A and hepatitis B virus IgM antibodies (a marker of acute infection) using ELISA (DiaSorin, Italy)

### Statistical analysis

All analyses were conducted separately for data from South Sudan and Bangladesh. We defined confirmed hepatitis E infection across sites as the presence of anti-HEV IgM antibodies in serum detected by ELISA. In primary analyses, we excluded suspected cases under the age of 14 from South Sudan since only individuals aged 14 and above were eligible for inclusion in Bangladesh. We excluded suspected cases with jaundice onset more than 30 days before the clinic visit/hospitalization in both study populations to focus on more typical conceptions of acute disease. In primary analyses patients were considered to have a symptom if they reported experiencing it at any point during their illness, even if it was not present on the day of their clinic visit/admission.

We first assessed the sensitivity (proportion of confirmed hepatitis E cases who satisfy a given case definition) and specificity (proportion of individuals without laboratory confirmed hepatitis E who do not satisfy a given case definition) of international case definitions for hepatitis E in the WHO hepatitis E outbreak toolbox (19) for identifying hepatitis E compared to laboratory confirmed hepatitis E and estimated 95% Wald confidence intervals. Laboratory confirmed hepatitis E was defined as the presence of anti-HEV IgM detected by ELISA given the higher sensitivity and specificity of ELISA relative to PCR and RDTs, high sensitivity of ELISA up to 30 days after jaundice onset (>90%) (22), and since PCR testing was not conducted for a majority of AJS patients in Bangladesh. The WHO recommended case definition for suspected hepatitis E during outbreaks is anyone with either or both of the following: (i) discrete onset of an acute illness with symptoms of acute infectious disease (fever, malaise, or fatigue) and signs of liver damage (anorexia, nausea, jaundice, dark urine, or right upper quadrant tenderness); and/or (ii) raised alanine aminotransferase (ALT) levels more than ten times the upper limit of normal laboratory levels (19). Given our focus on suspected case definitions, we limited our evaluation to the symptom-based clinical criteria outlined in the WHO case definition, excluding laboratory findings. We were unable to evaluate the full WHO suspected case definition in either setting since no data were collected on tenderness in the upper right quadrant. MSF defines suspected hepatitis E as the recent onset of jaundice plus the presence of one or more of the following: malaise, anorexia, epigastric discomfort, or nausea (9). International Medical Corps (IMC) defines suspected hepatitis E as clinically detected jaundice and one or more of the following: malaise, loss of appetite, fever, abdominal pain, or joint pain (9). In both study settings, fatigue/drowsiness was used interchangeably with malaise. Epigastric discomfort and joint pain were only collected in South Sudan and therefore, weren’t considered when evaluating the MSF or IMC case definitions in Bangladesh. A summary of signs and symptoms included in each international case definition for suspected hepatitis E is provided in Table 1 and signs and symptoms collected in each study population is provided in Table 2.

**Table 1.**
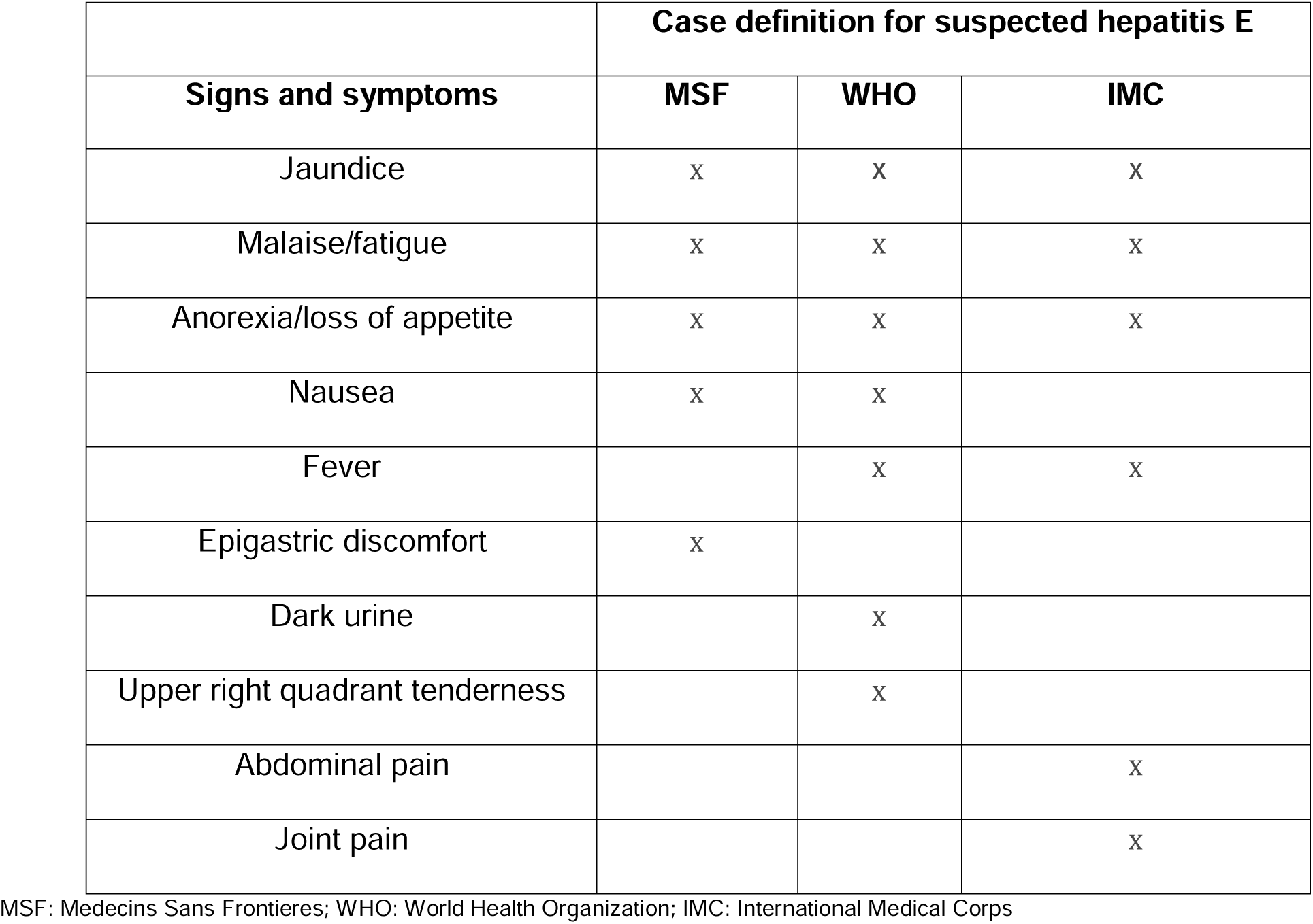
Summary of signs and symptoms in international case definitions for suspected hepatitis E in the World Health Organization hepatitis E outbreak toolbox. (**19**)

| Signs and symptoms | Case definition for suspected hepatitis E |  |  |
| --- | --- | --- | --- |
|  | MSF | WHO | IMC |
| Jaundice | X | X | X |
| Malaise/fatigue | X | X | X |
| Anorexia/loss of appetite | X | X | X |
| Nausea | X | X |  |
| Fever |  | X | X |
| Epigastric discomfort | X |  |  |
| Dark urine |  | X |  |
| Upper right quadrant tenderness |  | X |  |
| Abdominal pain |  |  | X |
| Joint pain |  |  | X |

MSF: Medecins Sans Frontieres; WHO: World Health Organization; IMC: International Medical Corps

**Table 2.** Summary of signs and symptoms collected in Bentiu, South Sudan and Bangladesh.

| Signs and symptoms | Bentiu | Bangladesh |
| --- | --- | --- |
| Yellow skin<sup>δ</sup> | X | X |
| Yellow eyes<sup>δ</sup> | X | X |
| Dark urine<sup>δ</sup> | X | X |
| Pale stools<sup>δ</sup> | X |  |
| Fever | X | X |
| Loss of appetite | X | X |
| Nausea | X | X |
| Diarrhoea | X | X |
| Vomiting | X | X |
| Headache | X | X |
| Abdominal pain | X | X |
| Altered mental state | X | X |
| Fatigue/drowsy | X | X |
| Convulsion | X | X |
| Malaise/fatigue | X | X |
| Anorexia/loss of appetite | X | X |
| Epigastric discomfort | X |  |
| Cough | X |  |
| Joint pain | X |  |
| Itch | X |  |
| Bleeding | X |  |
| Melaena |  | X |
| Constipation |  | X |
| Unconsciousness |  | X |
| Distended abdomen<sup>δδ</sup> |  | X |
| Oedema<sup>δδ</sup> |  | X |
| Dehydration<sup>δδ</sup> |  | X |
| Liver palpable<sup>δδ</sup> |  | X |

<sup>δ</sup>Signs and symptoms classically associated with acute jaundice syndrome
<sup>δδ</sup>Based on physician examination

To provide an effective “upper bound” on how well signs and symptoms, when combined in any manner, could distinguish between AJS (as defined in each setting) with and without HEV, we used an ensemble of flexible classification models. We fit and combined results from four models – random forest, a generalized additive model, extreme gradient boosting, and lasso – using the SuperLearner package in R. We explored separate models to classify HEV positive AJS cases based on (i) signs and symptoms alone; (ii) signs and symptoms plus age, sex, and days since jaundice onset. Both models were run separately in each study population. We used 10-fold cross-validation to create the optimal weighted average of the models (i.e., the ensemble-learner) with larger weights given to models with lower cross-validated risk. We used leave-one-out cross-validation and the average ensemble-learner to estimate the area under the receiver operator characteristic curve (AUC) in each study population. We quantified uncertainty in AUC values using the 2.5^th^ and 97.5^th^ percentiles of 2,000 bootstrap samples drawn with replacement from the observed and predicted outcomes. Given a lack of standardized guidance on what is considered “acute” jaundice, in sensitivity analyses, we fit the ensemble learning model to the subset of jaundiced patients with jaundice onset within 14 days of the clinic visit. In sensitivity analyses we repeated the ensemble models among acute jaundice patients in Bangladesh classifying patients as having a symptom only if the symptom was present on the day of their clinic admission.

To explore alternative case definitions, we estimated the sensitivity, specificity, positive and negative predictive values for all combinations of signs and symptoms in each study setting. To balance sensitivity and specificity, we only considered alternative case definitions that were informative (sensitivity + specificity > 1). We explored case definitions based on the presence of at least one symptom in addition to jaundice, as well as definitions requiring at least two symptoms, and selected the alternative case definition in each setting with the highest specificity. If multiple case definitions had the same specificity, we selected the case definition with the highest sensitivity. If sensitivity was also tied, we selected the case definition with the fewest signs/symptoms (e.g., itch/fatigue selected over itch/fatigue/nausea). We repeated analyses in a combined dataset including both study populations and explored alternative case definitions using combinations of signs and symptoms that were present in both datasets. We applied post-stratification weights to ensure each study population contributed 50% to the overall sample, regardless of the original study population size. In sensitivity analyses, we repeated analyses evaluating existing international case definitions for suspected hepatitis E and exploring alternative case definitions in children under the age of 14 in South Sudan. Code and the minimal dataset for all analyses are available at: https://github.com/HopkinsIDD/hev-case-defintion.

### Ethics

Ethical approval for the parent study in South Sudan was obtained from the South Sudan Ministry of Health Research Ethics Board (RERB-MOH # 54/27/09/2022) and MSF (MSF ERB #2167). Ethical approval for the parent study in Bangladesh was obtained from the institutional review board of the icddr,b (Protocol # PR-14060).

## Results

We identified 524 patients at least 14 years of age with acute jaundice in South Sudan (March to December 2022) and 1925 patients hospitalized with acute jaundice in Bangladesh (December 2014 to September 2017). We excluded 68 patients in South Sudan and 280 patients in Bangladesh missing jaundice onset date or with jaundice onset more than 30 days of clinic visit or hospital admission and excluded 35 patients in South Sudan and 20 patients in Bangladesh with incomplete diagnostic test results and/or symptom history, yielding a final study population of 421 patients with acute jaundice in South Sudan and 1625 in Bangladesh. Among patients with acute jaundice, 20% in South Sudan (83 of 421) and 38% in Bangladesh (611 of 1625) had IgM antibodies detected in serum by ELISA (Table 3). In both study populations, a higher proportion of male suspected cases had IgM antibodies detected compared to females (26% versus 13% in South Sudan, 40% versus 32% in Bangladesh). Patients with acute jaundice in South Sudan had shorter delays to care seeking at the surveillance facility compared to Bangladesh (median 4 days versus 14 days; Figure S1). Among patients with acute jaundice who were anti-HEV IgM negative at the time of their clinic visit, a substantially higher proportion had anti-HEV IgG detected by ELISA in South Sudan compared to Bangladesh (93% vs. 30%). In South Sudan, 9% of acute jaundice patients had malaria infection (39 of 421), 23% had evidence of acute or chronic hepatitis B infection (96 of 421), and 1% had hepatitis C infection (6 of 421) (Table S1). Malaria and hepatitis C testing was not conducted in Bangladesh, and 8% of acute jaundice patients had acute hepatitis A infection (136 of 1625) and 34% had evidence of acute or chronic hepatitis B infection (555 of 1625).

**Table 3.** Characteristics of patients with acute jaundice who presented to care at a health facility in Bentiu, South Sudan (March to December 2022) and were admitted to six tertiary hospitals in Bangladesh (December 2014 to September 2017)

| Characteristic<br>N (col %) | South Sudan |  |  | Bangladesh |  |  |
| --- | --- | --- | --- | --- | --- | --- |
|  | Overall<br>N=421 | Anti-HEV<br>IgM Negative<br>N=338 | Anti-HEV<br>IgM Positive<br>N=83 | Overall<br>N=1625 | Anti-HEV<br>IgM Negative<br>N=1014 | Anti-HEV<br>IgM Positive<br>N=611 |
| Female | 210 (50) | 182 (54) | 28 (34) | 508 (31) | 346 (34) | 162 (27) |
| Age, median (IQR) | 24.8<br>(19.2, 33.4) | 26.1<br>(20.1, 35.2) | 20.1<br>(16.6, 27.1) | 28.0<br>(20.0, 41.0) | 30.0<br>(22.0, 50.0) | 25.0<br>(20.0, 32.0) |
| Age |  |  |  |  |  |  |
| 14-19 | 123 (29) | 83 (25) | 40 (48) | 304 (19) | 186 (18) | 118 (19) |
| 20-39 | 234 (56) | 199 (59) | 35 (42) | 847 (52) | 439 (43) | 408 (67) |
| 40-59 | 53 (13) | 45 (13) | 8 (10) | 333 (20) | 254 (25) | 79 (13) |
| 60+ | 11 (3) | 11 (3) | 0 (0) | 141 (9) | 135 (13) | 6 (1) |
| Days since jaundice onset, median (IQR) | 4.0<br>(3.0, 9.0) | 4.0<br>(3.0, 9.0) | 5.0<br>(3.0, 9.0) | 14.0<br>(8.0, 20.0) | 14.0<br>(7.0, 20.0) | 15.0<br>(8.0, 20.0) |
| Anti-HEV IgG positive | 390 (93) | 308 (91) | 82 (99) | 909 (56) | 307 (30) | 602 (99) |
| Symptoms |  |  |  |  |  |  |
| Yellow skin<sup>δ</sup> | 50 (12) | 35 (10) | 15 (18) | 1622 (100) | 1011 (100) | 611 (100) |
| Yellow eyes<sup>δ</sup> | 272 (65) | 207 (61) | 65 (78) | 1625 (100) | 1014 (100) | 611 (100) |
| Dark urine<sup>δ</sup> | 367 (87) | 292 (86) | 75 (90) | 1612 (99) | 1002 (99) | 610 (100) |
| Pale stools<sup>δ</sup> | 0 (0) | 0 (0) | 0 (0) | - | - | - |
| Fever | 335 (80) | 272 (80) | 63 (76) | 659 (41) | 425 (42) | 234 (38) |
| Loss of appetite | 159 (38) | 121 (36) | 38 (46) | 1187 (73) | 745 (73) | 442 (72) |
| Nausea | 67 (16) | 40 (12) | 27 (33) | 759 (47) | 460 (45) | 299 (49) |
| Diarrhoea | 100 (24) | 87 (26) | 13 (16) | 65 (4) | 39 (4) | 26 (4) |
| Vomiting | 102 (24) | 73 (22) | 29 (35) | 336 (21) | 212 (21) | 124 (20) |
| Headache | 92 (22) | 79 (23) | 13 (16) | 377 (23) | 242 (24) | 135 (22) |
| Abdominal pain | 20 (5) | 16 (5) | 4 (5) | 737 (45) | 450 (44) | 287 (47) |
| Altered mental state | 25 (6) | 19 (6) | 6 (7) | 83 (5) | 69 (7) | 14 (2) |
| Fatigue/drowsy | 76 (18) | 45 (13) | 31 (37) | 565 (35) | 377 (37) | 188 (31) |
| Convulsion | 1 (0.2) | 1 (0.3) | 0 (0) | 38 (2) | 28 (3) | 10 (2) |
| Epigastric pain | 118 (28) | 88 (26) | 30 (36) | - | - | - |
| Cough | 49 (12) | 46 (14) | 3 (4) | - | - | - |
| Joint pains | 207 (49) | 165 (49) | 42 (51) | - | - | - |
| Itch | 10 (2) | 7 (2) | 3 (4) | - | - | - |
| Bleeding | 7 (2) | 6 (2) | 1 (1) | - | - | - |
| Melaena | - | - | - | 102 (6) | 67 (7) | 35 (6) |
| Constipation | - | - | - | 430 (26) | 225 (22) | 205 (34) |
| Unconsciousness | - | - | - | 50 (3) | 33 (3) | 17 (3) |
| Distended abdomen<sup>δδ</sup> | - | - | - | 225 (14) | 165 (16) | 60 (10) |
| Oedema<sup>δδ</sup> | - | - | - | 143 (9) | 119 (12) | 24 (4) |
| Dehydration<sup>δδ</sup> | - | - | - | 307 (19) | 184 (18) | 123 (20) |
| Liver palpable<sup>δδ</sup> | - | - | - | 177 (11) | 137(14) | 40 (7) |
| Hospitalized | 28 (7) | 22 (7) | 6 (7) | 1625 (100) | 1014 (100) | 611 (100) |
| Died* | 6 (1) | 5 (1) | 1 (1) | 51 (3) | 41 (4) | 10 (2) |

HEV: hepatitis E virus; IgM: immunoglobulin M; IQR: interquartile range
<sup>δ</sup>Signs and symptoms classically associated with acute jaundice syndrome
<sup>δδ</sup>Based on physician examination
\*Missing for 31 acute jaundice cases in Bangladesh (23 anti-HEV IgM negative, 8 anti-HEV IgM positive). Death during hospitalization in South Sudan and during hospitalization and up to 3 months after hospital discharge in Bangladesh.

### International case definitions for suspected hepatitis E

The sensitivity and specificity of existing international case definitions varied by study population (Table 4). The MSF case definition (AJS plus at least one of: fatigue/malaise, epigastric discomfort, nausea, anorexia/loss of appetite) had high sensitivity in both study populations (∼70-80%), but higher specificity in South Sudan (42%; 95% CI: 37, 47) compared to Bangladesh (17%; 95% CI: 15, 19). The IMC case definition (jaundice plus at least one of: fatigue/malaise, loss of appetite, fever, abdominal pain, joint pain) had the highest sensitivity and lowest specificity in both study populations. All international case definitions for suspected hepatitis E were misinformative in Bangladesh (sensitivity + specificity < 1), and the MSF case definition was the only informative case definition in South Sudan with specificity above 40%. The MSF and IMC case definitions had similar sensitivity and specificity in children under 14 years of age in South Sudan compared to older suspected cases, while the WHO case definition had higher sensitivity and lower specificity (Table S4).

**Table 4.**
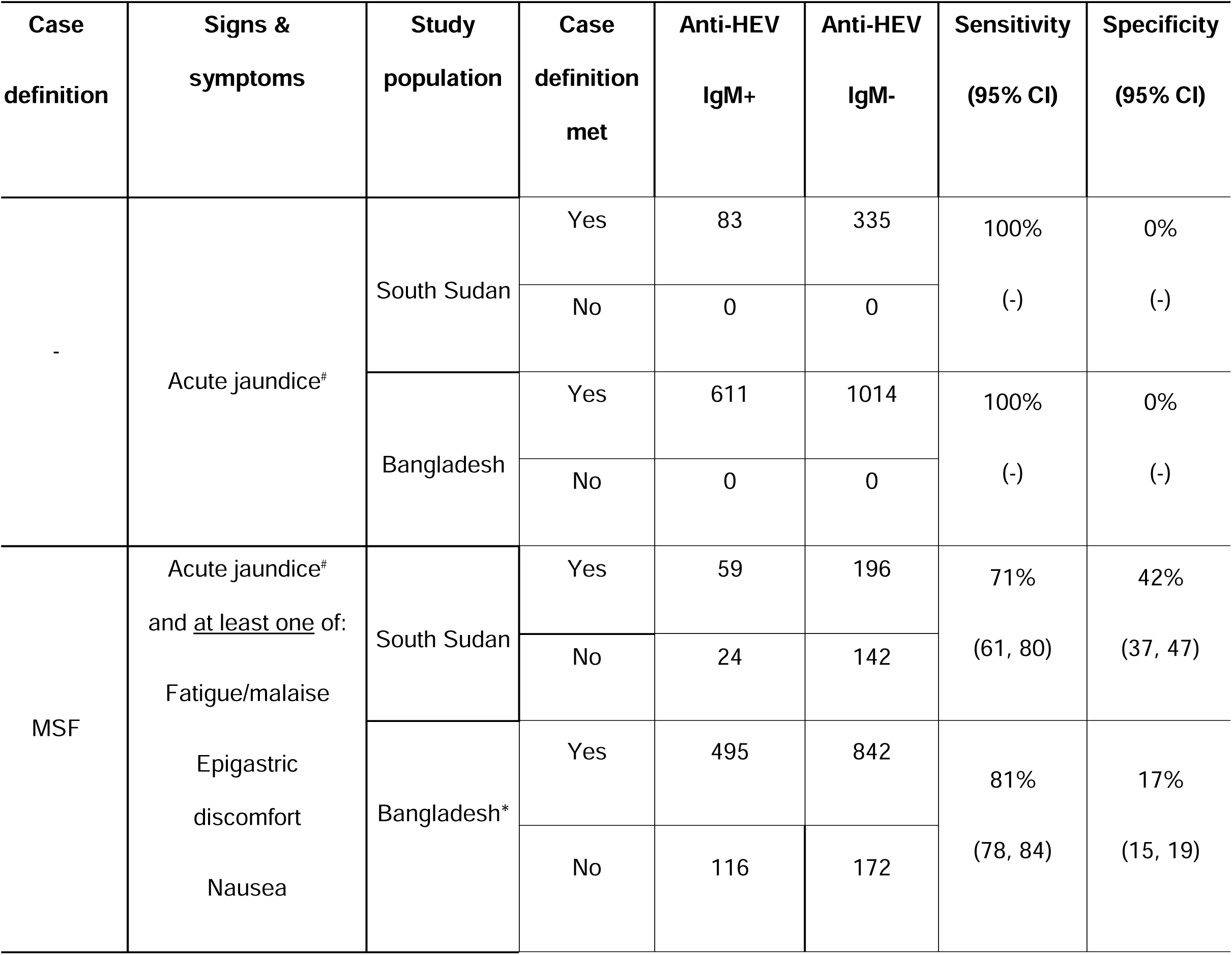
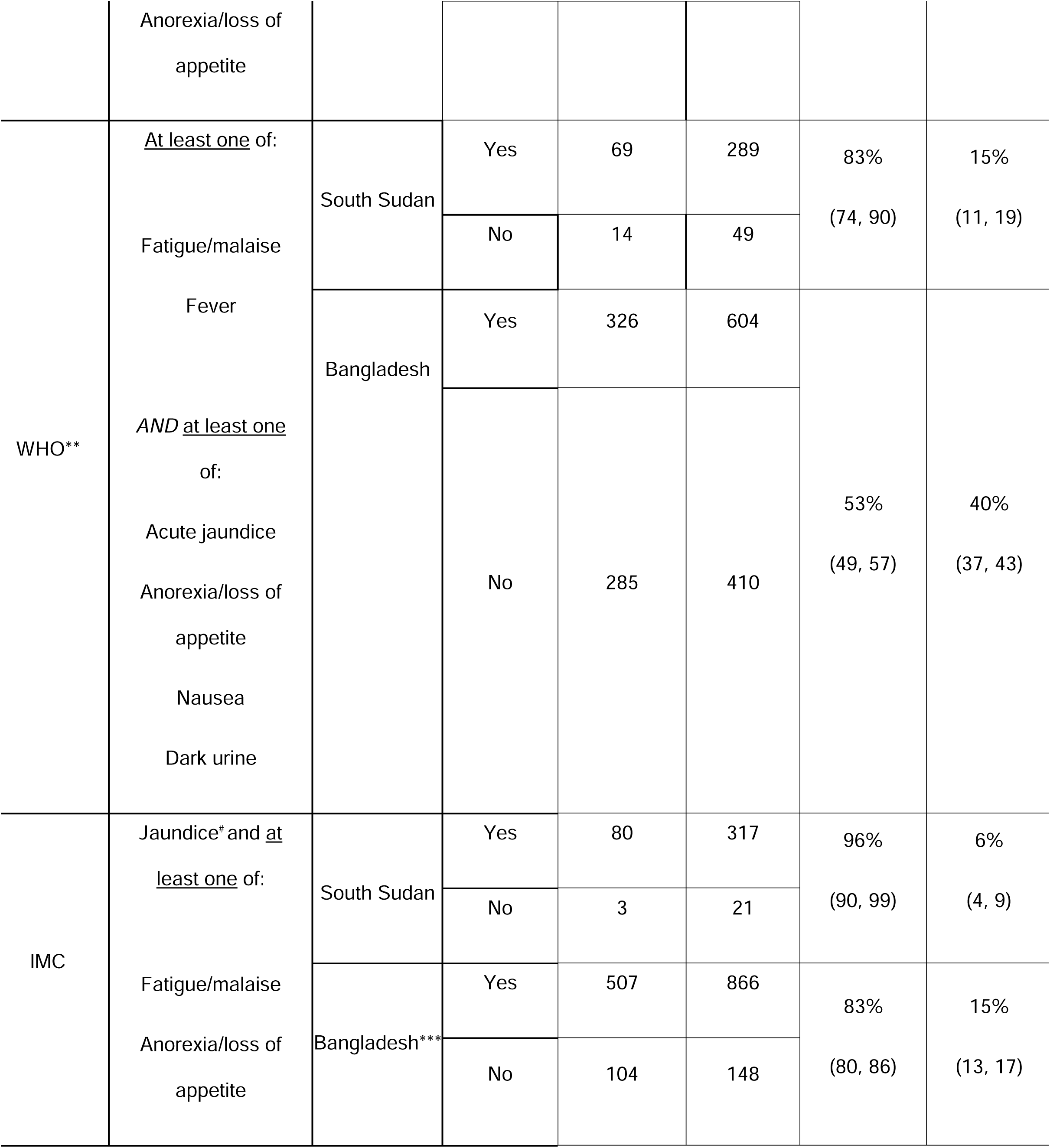
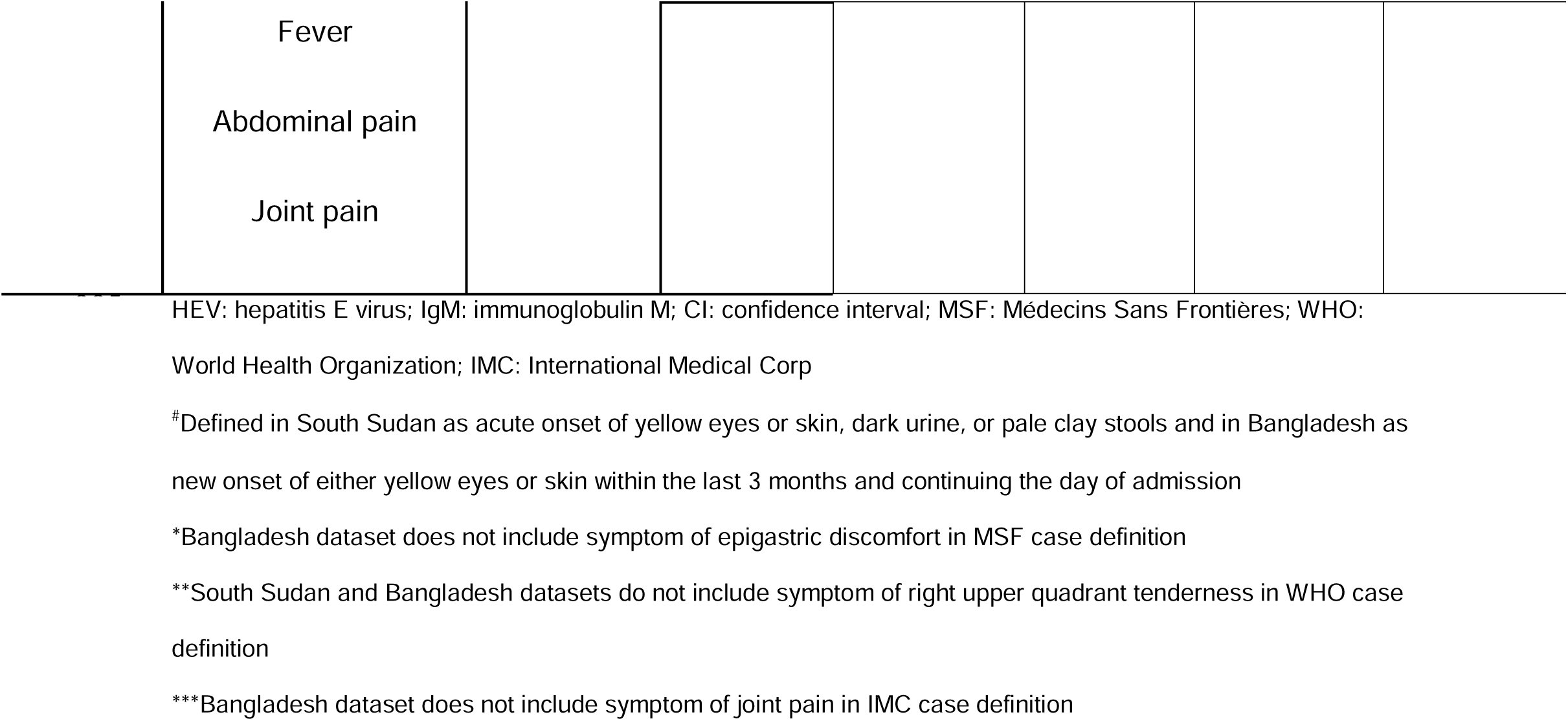
Sensitivity and specificity of international case definitions for suspected hepatitis E compared to laboratory-confirmed hepatitis E among patients with acute jaundice in South Sudan and Bangladesh.

### Alternative case definitions for suspected hepatitis E

Signs and symptoms alone had poor discrimination (AUC_SouthSudan_=0.64; 95% CI: 0.57, 0.71; AUC_Bangladesh_= 0.60; 95% CI: 0.57, 0.63) (Figure 1) in both study populations. Ensemble-learning models including age, sex, and days since jaundice onset had improved but moderate ability to discriminate between uninfected and infected suspected cases (AUC_SouthSudan_= 0.74; 95% CI: 0.68, 0.80; AUC_Bangladesh_= 0.73; 95% CI: 0.71, 0.76) relative to symptoms alone. Results were similar when restricting analyses to suspected cases with AJS onset within 14 days of the clinic visit (Figure S2). In sensitivity analyses discrimination was similar in patients with acute jaundice in Bangladesh based only on signs/symptoms at the time of hospital admission (AUC: 0.60; 95% CI: 0.57, 0.63) and signs and symptoms at the time of admission plus demographics (AUC=0.73; 95% CI: 0.71, 0.76) (Figure S3). Discrimination was lower in patients with acute jaundice under 14 years of age in South Sudan compared to older patients with acute jaundice for signs and symptoms alone (AUC=0.54; 95% CI: 0.48, 0.60) and signs and symptoms plus demographics (AUC=0.60; 95% CI: 0.54, 0.66) (Figure S4).

**Figure 1.**
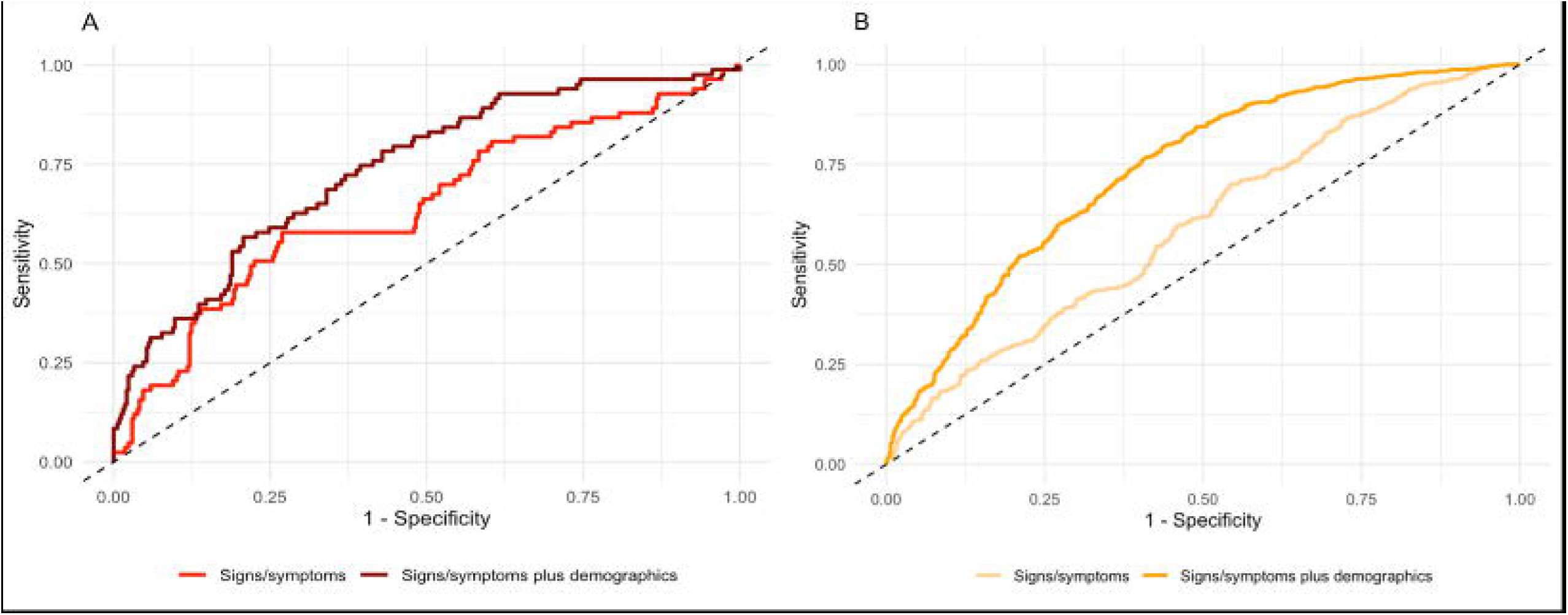
Receiver operator characteristic curves for ensemble-learning models predicting hepatitis E infection in South Sudan (A) and Bangladesh (B). Demographic variables include age, sex, and days since jaundice onset. Bleeding, convulsions, and itch were excluded from ensemble-learning models in South Sudan due to low prevalence. Dashed line indicates discrimination no better than chance alone.

Because ensemble models are impractical for real-world application and were intended only to represent an effective upper bound on potential performance, we next explored all combinations of signs/symptoms among patients with acute jaundice as alternative case definitions. Among the informative (sensitivity + specificity > 1) alternative case definitions with the highest specificity (Table 5), all had higher specificity relative to existing case definitions. All alternative case definitions had sensitivity and specificity below 70% in South Sudan and below 60% in Bangladesh. Signs and symptoms in alternative case definitions with the highest specificity differed in each setting. In analyses combining study populations, no alternative case definitions had both sensitivity and specificity higher than 60% but alternative case definitions including abdominal pain and fatigue/malaise had a moderately high positive predictive value (77%; 95% CI: 74, 80). Case definitions requiring at least two symptoms had similar sensitivity and specificity as case definitions requiring at least one symptom. Among children under 14 in South Sudan, all alternative case definitions had sensitivity and specificity below 65% and had lower positive predictive values (65-69%) compared to alternative case definitions in older acute jaundice patients in South Sudan (88%) (Table S5).

**Table 5.**
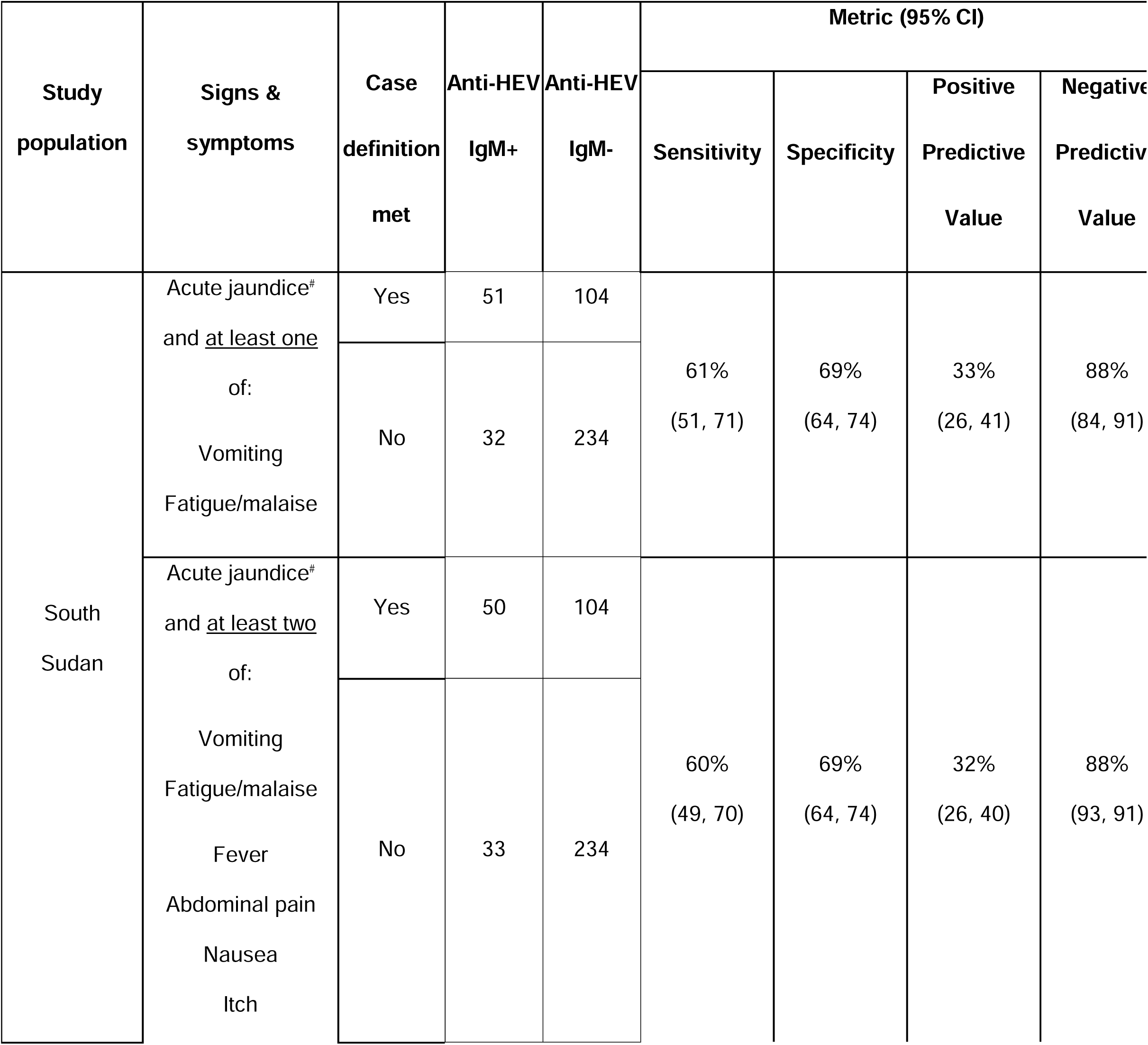
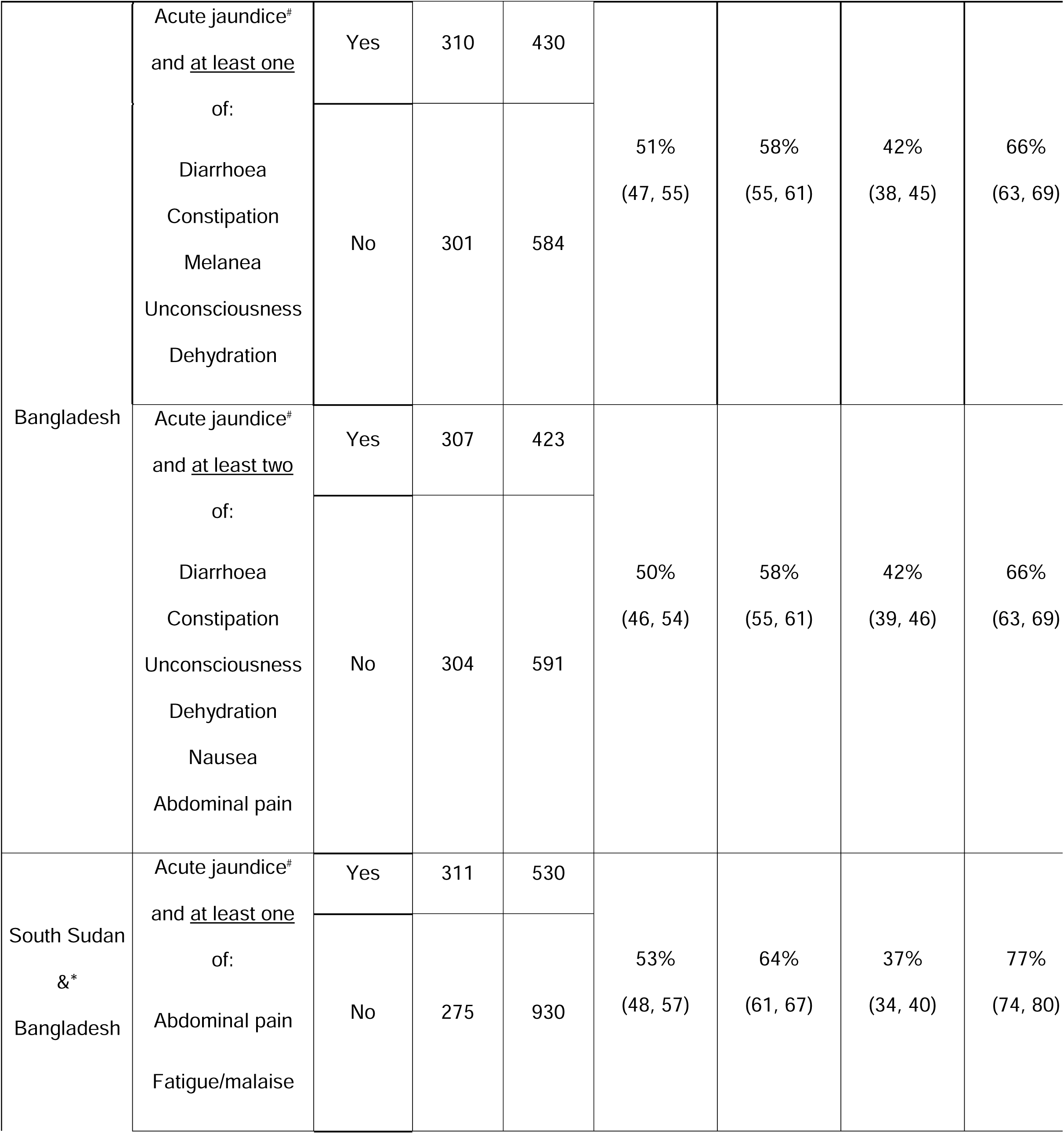
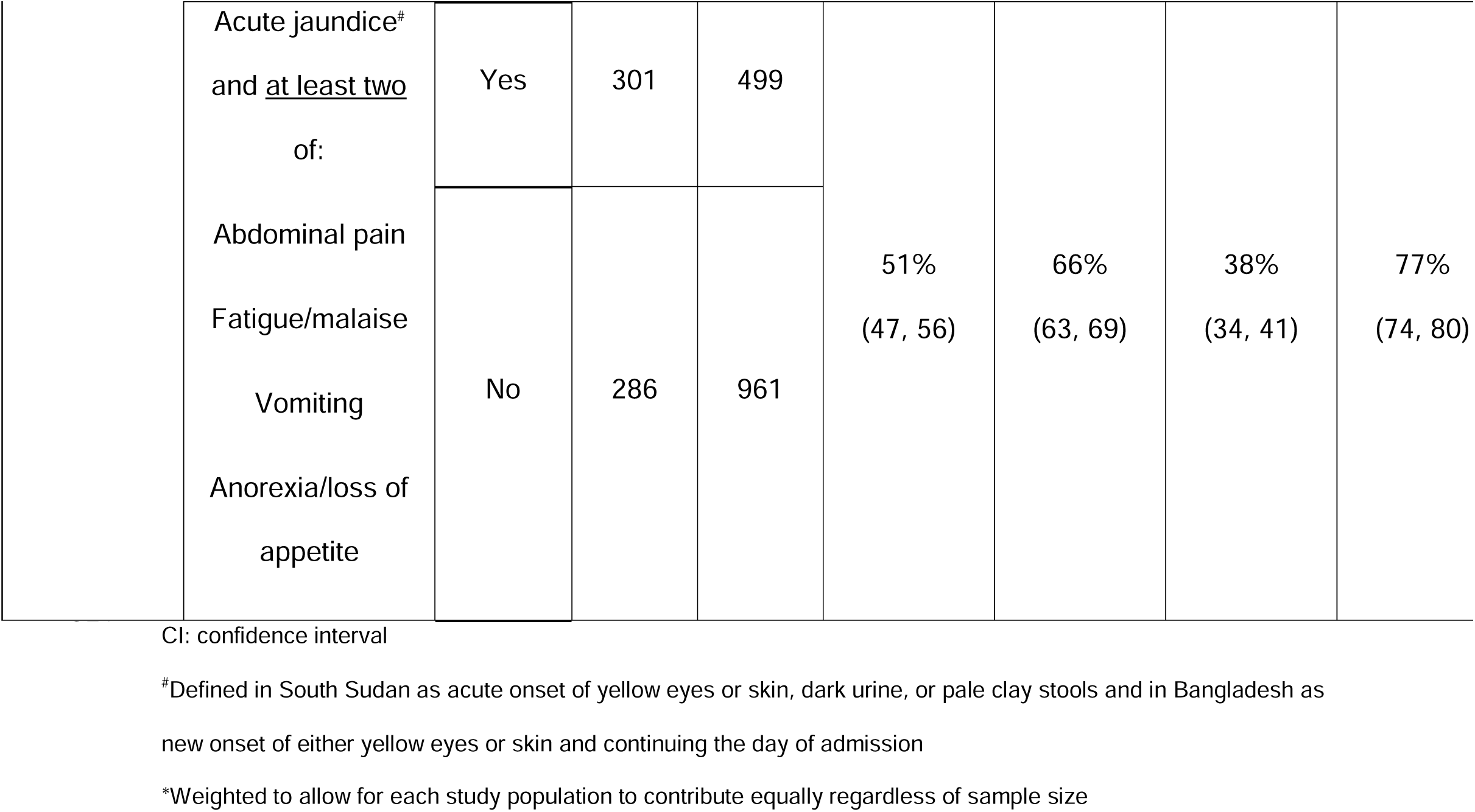
Sensitivity and specificity of informative alternative case definitions for suspected hepatitis E with the highest specificity compared to laboratory-confirmed hepatitis E in South Sudan and Bangladesh.

## Discussion

Case definitions serve an important role not only in the clinical management of patients, but in assuring that surveillance data are interpretable and can shed light on disease burden. Existing international case definitions for suspected hepatitis E recommended by the WHO and NGOs had variable sensitivity (53-96%) and specificity (6-60%) compared to laboratory-confirmed hepatitis E across populations of patients with acute jaundice syndrome (AJS). While most existing suspected case definitions identified a substantial proportion of true hepatitis E cases, their ability to correctly identify negatives was often worse than chance alone. Alternative case definitions for suspected hepatitis E, including information on a broad range of signs and symptoms and demographics, had suboptimal ability to discriminate between AJS with and without hepatitis E, and no signs/symptoms consistently improved specificity across study populations. Given the limitations of signs and symptoms, broader use of diagnostic tests for hepatitis E can facilitate a better understanding of the true burden of hepatitis E. In settings where use of ELISA and PCR for routine diagnosis of hepatitis E remains unlikely due to limited laboratory infrastructure and resources, expanding access to rapid diagnostic tests can help advocate for the timely use of existing interventions.

Similar studies evaluating case definitions for other syndromically identified infectious diseases have had variable results. Among suspected cholera cases with acute watery diarrhoea in surveillance data from seven countries in the African Cholera Surveillance Network, adding signs/symptoms to the existing suspected case definition of acute watery diarrhoea improved specificity without a substantial decrease in sensitivity (23). In contrast, for Zika virus disease, international suspected case definitions that included rash in facility-based surveillance in Singapore had much higher specificity than those that didn’t, but also had much lower sensitivity (24). Though no alternative case definitions had both sensitivity and specificity higher than 60% in our combined study population, multiple alternatives had higher specificity than the current international case definitions for suspected hepatitis E (Figure S5). These alternative case definitions for suspected hepatitis E could be considered for use in outbreak settings where rapid diagnostic tests (RDTs) are unavailable and where suspected case definitions with higher specificity may be preferred to inform outbreak response. The high negative predictive value of AJS plus abdominal pain or fatigue/malaise in our study population (77%, 95% CI: 74, 80), for example, could inform decision-making about whether to prioritize interventions best-suited to interrupt hepatitis E transmission (e.g. water and sanitation) over interventions better-suited for interrupting transmission of other pathogens. In contrast, the use of AJS alone as a case definition for suspected hepatitis E may be preferred in routine surveillance aimed at minimizing false negatives and understanding burden of disease or in settings with many competing causes of acute jaundice in which the prevalence of hepatitis E among acutely jaundiced patients may be lower (e.g. settings endemic for hepatitis E and other infectious causes of jaundice).

Our findings support recent recommendations from the Viral Hepatitis Group at the European Society of Clinical Microbiology and Infectious Diseases, emphasizing the importance of point-of-care testing platforms using RDTs for HEV detection, particularly in settings where laboratory capacity and infrastructure is not sufficient for routinely conducting ELISA or PCR. While our study population largely excluded pediatric cases, the poor ability of signs and symptoms to discriminate between AJS with and without HEV in children under 14 years of age in South Sudan highlights the importance of confirmatory testing also among children. In 2022, the WHO Strategic Advisory Group of Experts on In Vitro Diagnostics (SAGE IVD) conditionally added RDTs for HEV to WHO’s list of essential in vitro diagnostics, noting that RDTs can aid diagnosis and surveillance of hepatitis E in settings where PCR and ELISA are not accessible, though additional information on test performance is needed (14). Although point-of-care tests for other infectious causes of acute jaundice, such as hepatitis A and malaria, are cheaper and more widely available than HEV RDTs, we identified frequent co-infections of hepatitis E with other infectious causes of acute jaundice (Table S1) and illustrate how using diagnostic tests for other infectious diseases is not a viable strategy for ruling out hepatitis E infection in the absence of hepatitis E diagnostics. Current WHO recommendations for the recognition and investigation of hepatitis E outbreaks note that comprehensive testing for all suspected cases is not needed, but evidence-based guidance on the number of samples that should be tested and who should be prioritized for testing is lacking (9). Among the signs and symptoms evaluated in our study none were strong candidates for prioritizing acute jaundice patients for testing with RDTs. Our findings suggest that the use of signs and symptoms other than acute jaundice to determine who should be tested with RDTs would not meaningfully improve the positive predictive value of RDTs relative to acute jaundice alone. In settings where routine use of RDTs is not feasible for all acute jaundice patients, quantifying the positive predictive value of acute jaundice during endemic and outbreak periods by systematically testing a subset of patients with RDTs could help adjust and better interpret burden of disease estimates.

This analysis has several limitations. Our study populations differed in many ways, including different historical exposures to HEV, differences in the underlying incidence of other causes of jaundice, different definitions for acute jaundice syndrome (South Sudan: acute onset of yellow eyes or skin, dark urine, or pale clay stools; Bangladesh: new onset of either yellow eyes or skin within the past 3 months and continuing the day of admission), and most importantly, case severity. In in South Sudan most patients with AJS were outpatients (93%), whereas all patients in Bangladesh were hospitalized. The prevalence of some signs and symptoms differed between outpatients and inpatients in South Sudan (e.g. altered mental state, loss of appetite, vomiting; Table S2), although certain symptoms among South Sudan outpatients more closely resembled those in Bangladesh than those among South Sudan inpatients (e.g., altered mental status). A symptom combination that consistently improves specificity across settings may exist, but we may not have identified it because not all symptoms were collected in both populations. Misclassification of symptoms was likely non-differential because all patients in our study populations were acutely jaundiced and ill even if they were not infected with HEV, which could lead to underestimates of predictive accuracy. We were unable to evaluate the full WHO suspected case definition in either setting due to a lack of data on tenderness in the upper right quadrant. However, all individuals in our study population had acute jaundice, therefore, the inclusion of data on tenderness in the upper right quadrant would not change our estimates of the sensitivity and specificity of the WHO suspected case definition. We also could not evaluate the full MSF or IMC suspected case definitions in Bangladesh due to a lack of data on joint pain or epigastric pain. Including these symptoms would likely have increased sensitivity at the expense of specificity.

We used ELISA to detect anti-HEV IgM, which has high sensitivity and specificity (13,25) but could miss infections among individuals seeking care shortly after infection if their IgM antibodies have not yet developed (26). Longer care seeking delays in Bangladesh (median delay: 14 days) likely minimize the likelihood of false negative ELISA results, but false negatives in South Sudan (median delay: 4 days) could have artificially lowered estimates of sensitivity and specificity if misclassification occurred non-differentially by signs/symptoms. Nevertheless, the sensitivity of IgM ELISA among all AJS cases identified in the South Sudan surveillance site declined with increasing delays between jaundice onset and care seeking and was high among cases who sought care in the first 14 days since AJS onset (95%; Credible Interval: 93, 98), and only 4 AJS cases had detectable RNA but no detectable IgM based on ELISA (22).

## Conclusion

Existing international case definitions for suspected hepatitis E had variable sensitivity and low specificity for identifying hepatitis E in acute jaundice surveillance in South Sudan and Bangladesh. Our findings demonstrate that modifying signs and symptoms included in case definitions for suspected hepatitis E is unlikely to meaningfully enhance understanding of the true burden of hepatitis E. Accurately distinguishing between AJS with and without HEV, as is needed for improved estimates of burden and risk, will require expanded access and strategic use of diagnostic tests, including rapid diagnostic tests .

## Data Availability

Code and the minimal dataset for all analyses are available at: https://github.com/HopkinsIDD/hev-case-defintion.

## Acknowledgements

We thank all the MSF and icddr,b staff who collected these data and the participants in South Sudan and Bangladesh. We thank the diagnostic laboratories of the University Hospitals of Geneva (HUG) for help with serological testing.

## Author contributions

Conceptualization: AK, ASA, EG; Methodology: AK, ASA, EG, RCP, SPL; Formal analysis: AK, RCP; Investigation: KVA, RN, CA, RCP, AN, AA; Resources: KVA, RN; Data curation: KVA, CA, RCP, AN, KCB, SAS; Writing – Original Draft: AK, RCP; Writing – Review & Editing: RN, CA, EG, JW, MH, MA, EG, ESG, FBL, DB, MR, JR, IC, IE, ASA, RCP, AN, KCB, SAS, KKP, AA, SU, SPL; Supervision: ASA, ESG, RCP; Project administration: RN, KVA, CA, EG, MA, RCP, AN, KCB, KKP, SU.

## Supporting Information

**Table S1. Diagnostic test results for alternative infectious causes of jaundice for acute jaundice patients who presented to care at a health facility in Bentiu, South Sudan (March to December 2022) and were admitted to six tertiary hospitals in Bangladesh (December 2014 to September 2017).**

**Table S2**. **Comparison of signs and symptoms of hepatitis E in among acute jaundice patients over 14 years of age in South Sudan, the subset of acute jaundice patients over 14 years of age in South Sudan who were hospitalized, and all hospitalized acute jaundice patients in Bangladesh by careseeking behavior.** Includes only signs and symptoms that were collected in both study populations.

**Table S3. Characteristics of patients with acute jaundice <u>under 14 years of age</u> who presented to care at a health facility in Bentiu, South Sudan (March to December 2022)**

**Table S4. Sensitivity and specificity of international suspected case definitions for identifying confirmed hepatitis E among <u>children under 14 years</u> of age with acute jaundice in South Sudan**

**Table S5. Sensitivity and specificity of informative alternative suspected case definitions with the highest specificity for identifying confirmed hepatitis E in <u>children under</u> 14 South Sudan and Bangladesh**

**Figure S1. Distribution of days since acute jaundice syndrome (AJS) onset at time of clinic visit or hospital admission among suspected hepatitis E cases in South Sudan and Bangladesh.**

**Figure S2. Receiver operator characteristic curves for ensemble-learning models predicting hepatitis E infection <u>among patients with jaundice onset within 14 days of</u> <u>clinic visit</u> in South Sudan (A) and Bangladesh (B).** Bleeding, convulsions, and itch were excluded from ensemble-learning models in South Sudan due to low prevalence.

**Figure S3. Receiver operator characteristic curves for ensemble-learning models predicting hepatitis E infection in Bangladesh based on <u>signs and symptoms at the time</u> <u>of hospital admission</u>.**

**Figure S4. Receiver operator characteristic curves for ensemble-learning models predicting hepatitis E infection in <u>children under 14 years of age</u> in South Sudan.** Demographics include age, sex, and days since jaundice onset. Bleeding, convulsion, and itch were excluded from the ensemble-learning due to low prevalence. Dashed line indicates discrimination no better than chance alone.

**Figure S5. Sensitivity and specificity of alternative suspected case definitions among patients with acute jaundice who presented to care in South Sudan and Bangladesh.** The combined dataset was weighted to allow for each study population to contribute equally regardless of sample size. Red shading indicates alternative case definitions with specificity lower than the WHO and IMC case definitions in both study populations, blue shading indicates specificity greater than all international suspected case definitions in each separate study population.

