## Supplementary figures and images for "Beyond Acute Jaundice: refining case definitions for suspected hepatitis E in South Sudan and Bangladesh"

### Supplemental Figure 1

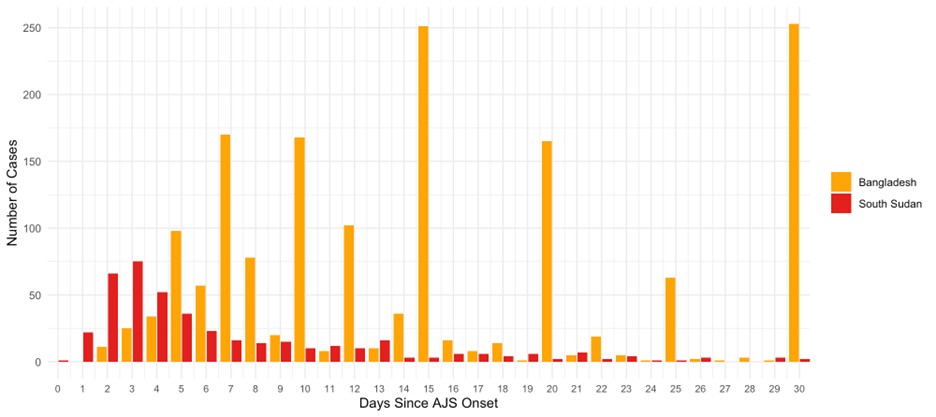

### Supplemental Figure 2

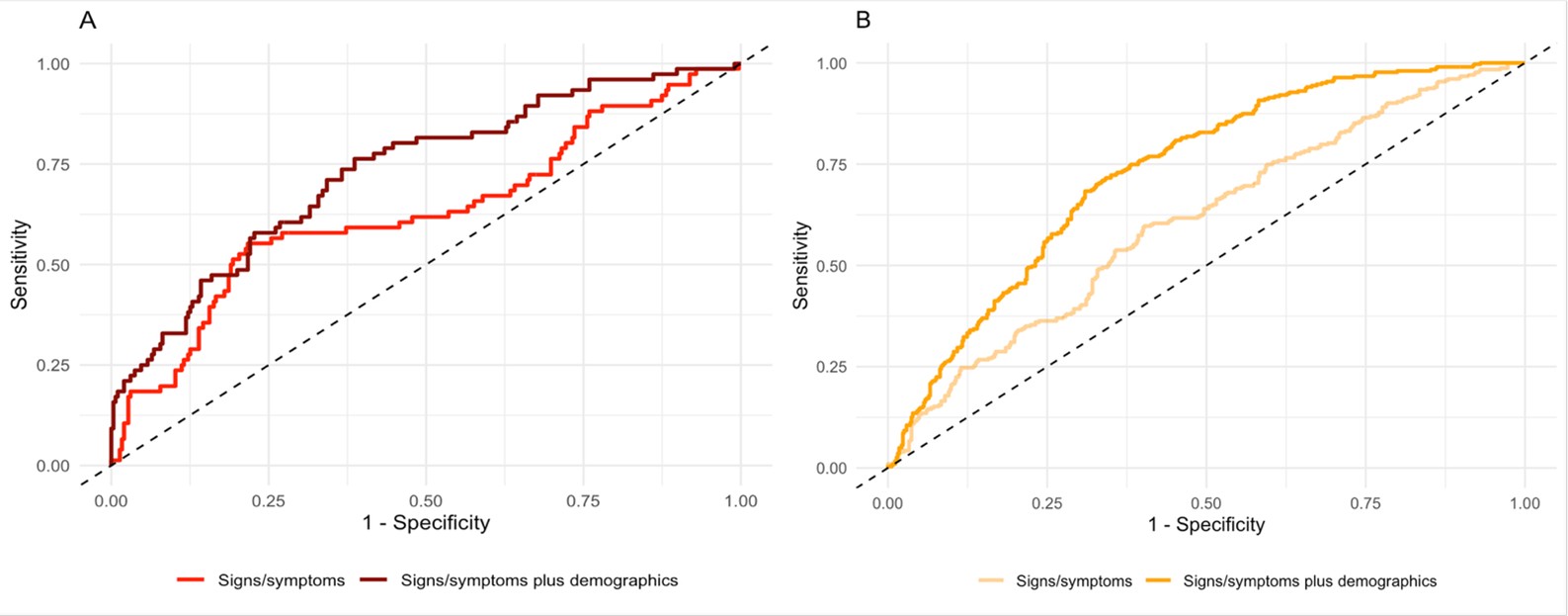

### Supplemental Figure 3

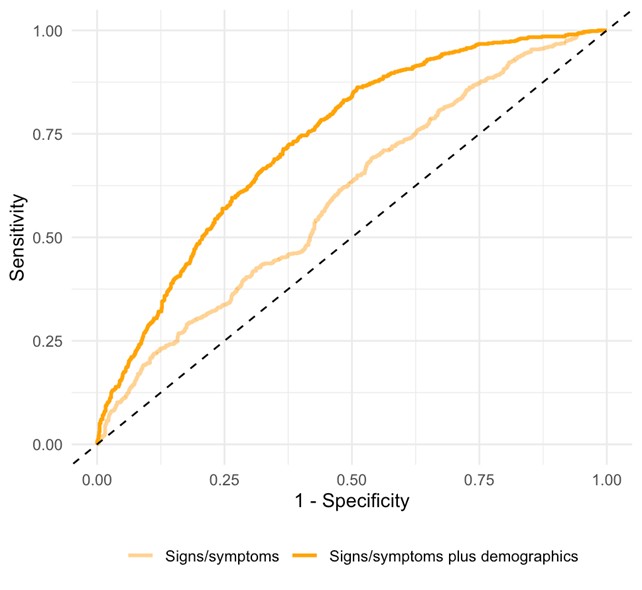

### Supplemental Figure 4

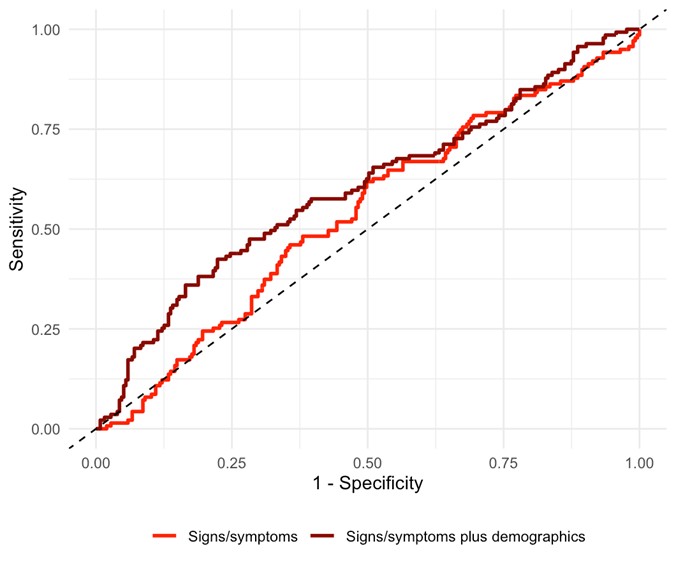

### Supplemental Figure 5

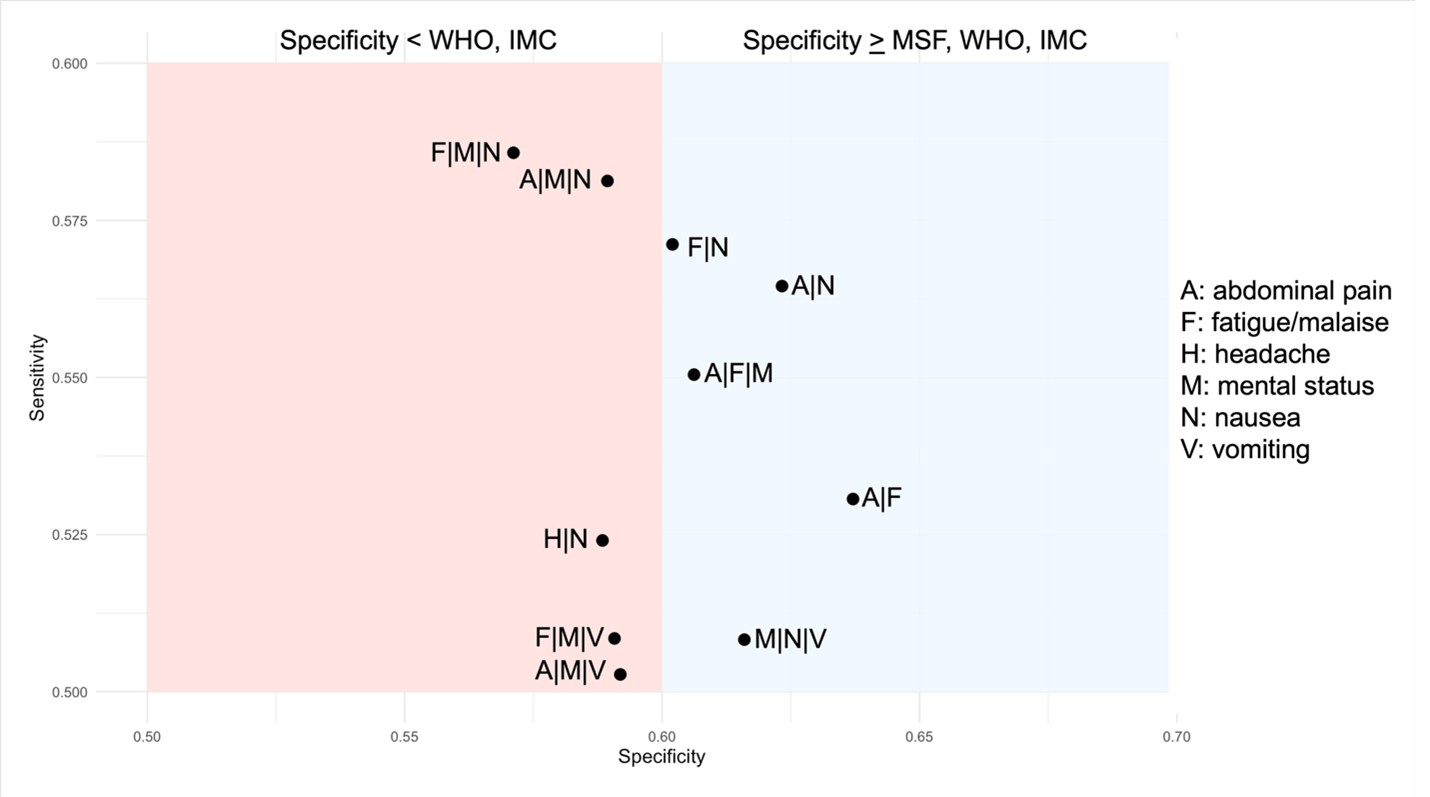
