## Supplemental Table 1 for "Beyond Acute Jaundice: refining case definitions for suspected hepatitis E in South Sudan and Bangladesh"

| **Characteristic**  **N (col %)** | **South Sudan** | | | **Bangladesh** | | |
| --- | --- | --- | --- | --- | --- | --- |
|  | **Overall**  N=421 | **Anti-HEV**  **IgM Negative**  N=338 | **Anti-HEV IgM Positive**  N=83 | **Overall**  N=1625 | **Anti-HEV**  **IgM Negative**  N=1014 | **Anti-HEV IgM Positive**  N=611 |
| Malaria infection* | 39 (9) | 36 (11) | 3 (4) | - | - | - |
| Hepatitis A infection | 0 (0) | 0 (0) | 0 (0) | 136 (8) | 128 (13) | 8 (1) |
| Hepatitis B infection** | 96 (23) | 83 (25) | 13 (16) | 555 (34) | 417 (41) | 138 (23) |
| Hepatitis C infection*** | 6 (1) | 5 (1) | 1 (1) | - | - | - |

HEV: hepatitis E virus; IgM: immunoglobulin M

*Missing for 1 acute jaundice patient in South Sudan (anti-HEV IgM negative)

**Missing for 10 acute jaundice patients cases in South Sudan (8 anti-HEV IgM negative, 2 anti-HEV IgM positive)

***Missing for 1 acute jaundice patient in South Sudan (anti-HEV IgM negative)
