## Supplemental Table 2 for "Beyond Acute Jaundice: refining case definitions for suspected hepatitis E in South Sudan and Bangladesh"

| Symptoms  (col%) | **Weeks between jaundice onset and clinic visit** | | | | | | | | |
| --- | --- | --- | --- | --- | --- | --- | --- | --- | --- |
|  | **≤ 1 week** | | | **>1 week to 2 weeks** | | | **>2 weeks to 1 month** | | |
|  | **SSD** | **SSD*** | **BD** | **SSD** | **SSD*** | **BD** | **SSD** | **SSD*** | **BD** |
| Fever | 82% | 73% | 42% | 76% | 75% | 39% | 72% | 100% | 42% |
| Loss of appetite | 37% | 50% | 76% | 44% | 50% | 70% | 34% | 0% | 73% |
| Nausea | 13% | 32% | 50% | 24% | 0% | 44% | 22% | 0% | 47% |
| Diarrhea | 26% | 41% | 6% | 15% | 25% | 3% | 22% | 100% | 4% |
| Vomiting | 23% | 73% | 26% | 28% | 50% | 22% | 24% | 50% | 17% |
| Headache | 20% | 5% | 23% | 23% | 25% | 23% | 32% | 50% | 23% |
| Abdominal pain | 5% | 0% | 42% | 4% | 0% | 45% | 10% | 0% | 47% |
| Altered mental state | 8% | 36% | 4% | 4% | 50% | 3% | 0% | 0% | 7% |
| Fatigue/drowsiness | 16% | 18% | 33% | 23% | 0% | 32% | 24% | 0% | 37% |
| Convulsion | 0% | 0% | 3% | 0% | 0% | 2% | 2% | 0% | 2% |

SSD: South Sudan; BD: Bangladesh

*Subset of acute jaundice patients over 14 years of age in South Sudan that were hospitalized (N=28)
