## Supplemental Table 3 for "Beyond Acute Jaundice: refining case definitions for suspected hepatitis E in South Sudan and Bangladesh"

| **Characteristic**  **N (col %)** | **South Sudan** | | | |
| --- | --- | --- | --- | --- |
|  | **Overall**  N=394 | | **Anti-HEV IgM Negative**  N=255 | **Anti-HEV IgM Positive**  N=139 |
| Female | 160 (41) | 88 (35) | | 72 (52) |
| Age, median (IQR) | 5.5 (3.4, 9.5) | 5.4 (3.2, 10.1) | | 5.8 (3.8, 8.0) |
| Age |  |  | |  |
| 0-4 | 160 (41) | 109 (43) | | 51 (37) |
| 5-10 | 70 (18) | 52 (20) | | 18 (13) |
| 11-13 | 164 (42) | 94 (37) | | 70 (50) |
| Days since jaundice onset,  median (IQR) | 4.0 (3.0, 7.0) | 4.0 (2.0, 7.0) | | 4.0 (3.0, 6.0) |
| Days since jaundice onset |  |  | |  |
| ≤1 week | 320 (81) | 206 (81) | | 114 (82) |
| >1 week to 2 weeks | 51 (13) | 30 (12) | | 21 (15) |
| >2 weeks to 1 month | 23 (6) | 19 (7) | | 4 (3) |
| Anti-HEV IgG positive | 307 (78) | 170 (67) | | 137 (99) |
| Symptoms |  |  | |  |
| Yellow skin^δ^ | 51 (13) | 29 (11) | | 22 (16) |
| Yellow eyes^δ^ | 320 (81) | 206 (81) | | 114 (82) |
| Dark urine^δ^ | 327 (83) | 202 (79) | | 125 (90) |
| Pale stools^δ^ | 16 (4) | 13 (5) | | 3 (2) |
| Fever | 349 (89) | 225 (88) | | 124 (89) |
| Loss of appetite | 208 (53) | 127 (50) | | 81 (58) |
| Nausea | 39 (10) | 27 (11) | | 12 (9) |
| Diarrhoea | 137 (35) | 93 (36) | | 44 (32) |
| Vomiting | 128 (32) | 85 (33) | | 43 (31) |
| Headache | 27 (7) | 16 (6) | | 11 (8) |
| Abdominal pain | 6 (2) | 4 (2) | | 2 (1) |
| Altered mental state | 20 (5) | 17 (7) | | 3 (2) |
| Fatigue/drowsy | 50 (13) | 33 (13) | | 17 (12) |
| Convulsion | 6 (2) | 6 (2) | | 0 (0) |
| Epigastric pain | 65 (16) | 50 (20) | | 15 (11) |
| Cough | 114 (29) | 80 (31) | | 34 (24) |
| Joint pains | 89 (23) | 61 (24) | | 28 (20) |
| Itch | 4 (1) | 4 (2) | | 0 (0) |
| Bleeding | 5 (1) |  | |  |
| Melaena | - | - | | - |
| Constipation | - | - | | - |
| Unconsciousness | - | - | | - |
| Distended abdomen^δ^ | - | - | | - |
| Oedema^δ^ | - | - | | - |
| Dehydration^δ^ | - | - | | - |
| Liver palpable^δ^ | - | - | | - |
| Hospitalized* | 25 (6) | 20 (8) | | 5 (4) |
| Died | 3 (1) | 3 (1) | | 0 (0) |

^δ^Signs and symptoms classically associated with acute jaundice syndrome

^δ^Based on physician examination

*Missing for 1 suspected case (IgM negative)
