## Supplemental Table 4 for "Beyond Acute Jaundice: refining case definitions for suspected hepatitis E in South Sudan and Bangladesh"

| **Study population** | **Case**  **definition** | **Signs & symptoms** |  | **Anti-HEV**  **IgM+** | **Anti-HEV**  **IgM-** | **Sensitivity**  **(95% CI)** | **Specificity**  **(95% CI)** |
| --- | --- | --- | --- | --- | --- | --- | --- |
| South  Sudan | MSF* | Acute jaundice^#^  and at least one of:  Fatigue/malaise  Epigastric discomfort  Nausea  Anorexia/loss of appetite | Yes | 95 | 162 | 68%  (60, 75) | 36%  (31, 43) |
|  |  |  | No | 44 | 93 |  |  |
|  | WHO* | At least one of:  Fatigue/malaise  Fever  *AND* at least one of:  Acute jaundice  Anorexia/loss of appetite  Nausea  Dark urine | Yes | 127 | 229 | 91%  (86, 95) | 10%  (7, 15) |
|  |  |  | No | 12 | 26 |  |  |
|  | IMC | Jaundice^#^ and at least one of:  Fatigue/malaise  Anorexia/loss of appetite  Fever  Abdominal pain  Joint pain | Yes | 135 | 245 | 97%  (93, 99) | 4%  (2, 7) |
|  |  |  | No | 4 | 10 |  |  |

HEV: hepatitis E virus; IgM: immunoglobulin M; CI: confidence interval; MSF: Médecins Sans Frontières; WHO: World Health Organization; IMC: International Medical Corp

^#^Defined in South Sudan as acute onset of yellow eyes or skin, dark urine, or pale clay stools and in Bangladesh as new onset of either yellow eyes or skin continuing the day of admission

*Dataset does not include symptom of right upper quadrant tenderness in WHO case definition
