## Supplemental Table 5 for "Beyond Acute Jaundice: refining case definitions for suspected hepatitis E in South Sudan and Bangladesh"

| **Study population** | **Signs & symptoms** | **Case**  **definition**  **met** | **Anti-HEV**  **IgM+** | **Anti-HEV**  **IgM-** | **Metric (95% CI)** | | | |
| --- | --- | --- | --- | --- | --- | --- | --- | --- |
|  |  |  |  |  | **Sensitivity** | **Specificity** | **Positive**  **Predictive**  **Value** | **Negative**  **Predictive**  **Value** |
| South Sudan | Acute jaundice^#^  and at least one of:  Vomiting  Abdominal pain  Headache | Yes | 56 | 99 | 40%  (33, 49) | 61%  (55, 67) | 36%  (29, 44) | 65%  (59, 71) |
|  |  | No | 83 | 156 |  |  |  |  |
|  | Acute jaundice^#^  and at least two of:  Fever  Loss of appetite | Yes | 74 | 111 | 54%  (46, 62) | 56%  (50, 62) | 40% (33, 47) | 69%  (62, 75) |
|  |  | No | 64 | 137 |  |  |  |  |

CI: confidence interval

^#^Defined in South Sudan as acute onset of yellow eyes or skin, dark urine, or pale clay stools
